# The dynamically neurodiverse human brain: Measuring excitatory-inhibitory dynamics and identifying homeostatic differences in autistic and non-autistic people

**DOI:** 10.1101/2023.06.19.23291507

**Authors:** C. L. Ellis, J. Ahmad, A. Zoumpoulaki, M. Dimitrov, H. E. Velthuis, A. C. Pereira, N. M. L. Wong, M. F. Ponteduro, L. Kowalewski, A. Leonard, P. Garces, Q. Huang, E. Daly, D. Murphy, G. McAlonan

## Abstract

Brain function is the dynamic output of coordinated excitatory and inhibitory (E-I) activity. E-I alterations, arising from differences in excitatory glutamate and inhibitory GABA pathways, are implicated in the development and heterogeneity of multiple neurodevelopmental conditions, such as autism; and are consequently targets for pharmacological support options. Yet, E-I measures of neurotransmitter levels or receptors in the living human brain (such as Magnetic Resonance Spectroscopy or Positron Emission Tomography) are expensive and/or invasive and do not capture dynamics. The determine if a candidate metric captures a neurosignalling system, the system must be challenged and changes observed objectively. This is basis of animal study designs. The aperiodic 1/f exponent of the EEG power spectrum is sensitive to E-I perturbations in animals but, more work is needed to translate to humans. Therefore, we tested the hypotheses that i) the aperiodic 1/f exponent of resting-state EEG in humans changes following a pharmacological E-I challenge with arbaclofen, a GABA_B_ receptor agonist; and ii) dynamic responsivity to GABAergic challenge is different in a neurodevelopmental condition associated with E-I differences, namely autism. As predicted, in both groups the aperiodic 1/f exponent significantly increased following a high (30mg) dose of arbaclofen. However, an aperiodic exponent increase was also elicited at a lower (15mg) dose of arbaclofen in autistic but not non-autistic individuals. Hence, in humans, the aperiodic 1/f exponent captures E-I dynamics and autistic brains are dynamically different compared to non-autistic brains. We suggest that our results can be explained by homeostatic differences E-I regulation between groups.

## Introduction

Excitatory and inhibitory (E-I) signalling is a fundamental property of brain. Whilst it involves the interplay of numerous chemical transmitter systems, glutamate is the most abundant excitatory neurotransmitter in the postnatal human brain, whereas GABA is the primary inhibitory neurotransmitter. A tightly organised E-I relationship emerges during early development (Dorrn et al., 2010), adapts to brain state (i.e. rest(Okun & Lampl, 2008) and sleep-stages (Niethard et al., 2016)) and is crucial for efficient cortical functioning. For example, animal studies have shown how, at the synaptic level, dynamic E-I signalling serves to refine and maintain precisely-tuned responses to stimuli across multiple sensory domains (Heiss et al., 2008; Mariño et al., 2005; Poo & Isaacson, 2009; Tao & Poo, 2005; Wehr & Zador, 2003), as well as higher cognitive functions such as memory (Lim & Goldman, 2013; Rubin et al., 2017; Vogels et al., 2011), information processing and social behaviour (Yizhar et al., 2011).

Alterations in the regulation of E-I signalling have also been implicated in multiple neurodevelopmental conditions which impact upon sensory processing, sleep and higher order cognition, such as autism (Sohal & Rubenstein, 2019) and schizophrenia (Kehrer et al., 2008). Indeed, genes and environmental exposures which increase the likelihood of having a neurodevelopmental condition, are known to change components of the E-I pathway (Gao & Penzes, 2016; Toro et al., 2010). For instance, autism has been associated with changes in genes that encode cell adhesion molecules, NRXNs and NLGNs; these are responsible for E and I synapse generation, specificity and function (Cao & Tabuchi, 2017; Südhof, 2008). These synapse level perturbations are subsequently thought to contribute to developmental circuit and network differences in the autistic brain (Toro et al., 2010).

However, despite consensus that E-I differences are associated with atypical neurodevelopment, we have few ways of assessing it in action in the living human brain. This is because, first, E-I signalling is complex: it is a dynamic process (Bridi et al., 2020; Brunwasser & Hengen, 2020) that exists at multiple interacting levels (single neuron, local populations of neurons or whole brain) - for example, local E-I changes after stroke interrupt global, whole-brain communication (Páscoa dos Santos & Verschure, 2022). Second, tools to investigate E-I processes in humans are limited. Current methods such as Magnetic Resonance Spectroscopy (MRS) and Positron Emission Tomography (PET) are expensive and/or invasive and, critically, lack the temporal resolution necessary to examine E-I dynamics (Lystad & Pollard, 2009; Pfister et al., 2014). Moreover, *changes* in E-I temporal dynamics can exist without altering static (or average) E-I properties (Bruining et al., 2020; Sohal & Rubenstein, 2019; Szücs & Huerta, 2015). With a temporal resolution well-suited to investigating E-I dynamics at faster timescales, EEG is a relatively inexpensive and safe tool which could help capture E-I brain dynamics in humans.

Specifically, the aperiodic component of the EEG power spectrum density (PSD) has been assumed to be a proxy of E-I signalling (Gao, Peterson, Voytek, et al., 2017; He et al., 2019; Molina et al., 2020; Peterson et al., 2017; Tran et al., 2020; Veerakumar et al., 2019). In the EEG PSD, the distribution of signal power over frequency follows a 1/f-like power law. The aperiodic component of the PSD represents non-oscillatory activity present in the absence of prominent oscillations (Donoghue, Dominguez, et al., 2020; Donoghue et al., 2021; Donoghue, Haller, et al., 2020), it can be modelled by a 1/f χ function (Donoghue, Haller, et al., 2020). The χ parameter, hereby referred to as the aperiodic 1/f exponent, is therefore used to quantify aperiodic activity and is equivalent to the slope of the EEG PSD (a.k.a. aperiodic slope). Gao et al. demonstrated that aperiodic 1/f exponent (Gao, Peterson, Voytek, et al., 2017), when captured from Local Field Potential and Electrocorticography recordings in rats and macaques, correlate with E (AMPA receptor positive): I (GABA_A_ receptor positive) synapse density ratio, vary dynamically with hippocampal theta oscillations, and steepen following computational simulations and pharmacological challenges which increase inhibition (Gao, Peterson, & Voytek, 2017).

However, the evidence that the aperiodic signal captures E-I relevant information in humans is less direct. Observations from a small sample of neurotypical participants indicate that aperiodic 1/f exponents are altered by the anaesthetics, ketamine and propofol (Waschke et al., 2021). Nonetheless, although ketamine and propofol act on glutamate and GABA receptors, respectively, with broad consequences for network activity (Taub et al., 2013) their action is non-specific; both interact with several other (non-E-I) receptors and ion channels (Eckenhoff & Tang, 2018; Zanos et al., 2018), limiting the interpretation of their effect. Furthermore, anaesthesia alters consciousness state; which is a potential confound because aperiodic 1/f exponents are known to vary with brain state from wakefulness to NREM to REM sleep (Lendner et al., 2020).

Additional evidence for the relationship between E-I and aperiodic 1/f exponent comes from a study of the effects of memantine (an NMDA, 5HT3 and Nicotinic acetylcholine receptor antagonist) on the aperiodic 1/f exponent in individuals with schizophrenia, a neuropsychiatric disorder with hypothesised E-I differences (Molina et al., 2020). Here, memantine was reported to ‘normalize’ baseline differences in aperiodic 1/f exponent in people with schizophrenia, but did not alter 1/f exponents in neurotypical people. An alternative explanation is that homeostatic E-I processes in the typical brain maintain the aperiodic slope at the dose of memantine administered in neurotypicals; whereas, the shift in aperiodic 1/f exponents in participants with schizophrenia reflects dynamic differences in homeostatic regulation of E-I in this neurodevelopmental condition. This offers the aperiodic 1/f exponent as a potentially useful metric to capture homeostatic E-I dynamics in the living human brain. However, constraining this interpretation is that aperiodic 1/f exponents were extracted from EEG data recorded during an auditory oddball task. Because aperiodic 1/f exponents are sensitive to auditory stimuli (Gyurkovics et al., 2022), and individuals with schizophrenia demonstrate atypical EEG responses to auditory oddball stimuli (Kaur et al., 2019; Lee et al., 2017; Light & Swerdlow, 2015; Salisbury et al., 2002), an atypical response to drug cannot be separated from an atypical response during task.

This Proof of Concept (PoC) study was designed to overcome a number of previous constraints in the use of the 1/f metric as a measure of E-I and potentially E-I homeostasis. First, a GABA_B_ receptor agonist, arbaclofen, was used to directly increase global inhibition in the human brain (de Groot & van Strien, 2018), without altering consciousness state, in adults with and without an E-I dependent neurodevelopmental condition (Autism Spectrum Disorder; autism) (Sohal & Rubenstein, 2019). Second, aperiodic 1/f exponents were extracted from EEG recorded at rest to eliminate the influence of task on aperiodic 1/f exponents. We hypothesized that increasing inhibition with arbaclofen would cause larger (steeper) aperiodic 1/f exponents compared to placebo, in line with the animal findings (Gao, Peterson, Voytek, et al., 2017). We also predicted that the dose-dependent change in 1/f slope would be different in autistic and non-autistic people.

## Materials and Method

### Design, participants, procedure

40 participants (25 non-autistic and 15 autistic) took part in this double-blind, placebo-controlled, repeated measures study. Participants were invited to take part in 3 study visits whereby they were given a single dose of arbaclofen (STX209; low dose = 15 mg, high dose = 30 mg) or a placebo, the order of which was randomised. 104 study visits were completed. EEG data were collected 3 hours post-drug and within the half-life of arbaclofen (Berry-Kravis et al., 2017). A medic was present for all study visits and monitored participants regularly for adverse side effects. At their discretion, in response to potential side effects, medics were then (and only then) able to access unblinding information. We noted that participants were more likely to experience known side effects of arbaclofen (nausea and dizziness) at the higher dose as expected, therefore our ethics committee approved an amendment to ensure that the order of administration was adjusted so that the high dose of arbaclofen was always after the low dose. This way participants who experienced particularly uncomfortable side effects could potentially avoid exposure to the higher dose of arbaclofen at a later visit.

Details regarding recruitment and diagnostic screening procedures have been provided previously (Huang et al., 2022). Briefly, autistic participants were either recruited from National Autism and ADHD Service for Adults (NAASA) at the South London and Maudsley National Health Service (NHS) Foundation Trust where diagnosis is a clinical decision supported by information from the Autism Diagnostic Interview–Revised where an informant is available and/or Autism Diagnostic Observation Schedule of current features. Where diagnosis information was provided from another clinic, their diagnostic process was reviewed by an experienced clinician at the screening interview. Participants gave informed consent, according to the Declaration of Helsinki, for a protocol as approved by the King’s College London Ethics Committee (Institutional Review Board). Exclusion criteria included IQ < 70, known autism-related genetic syndromes (e.g. fragile X syndrome or 22q11 deletion syndrome), medications directly affecting GABA or glutamate, significant comorbid psychiatric illness, epilepsy, known allergies to medication components and MRI-related contraindications.

Demographic data including age, biological sex, full-scale intelligence quotient and autism quotient are provided in table 1. Biological sex and full-scale IQ did not significantly differ between groups. As expected, there was a significant difference in autistic traits (AQ scores) between the autistic and non-autistic group. All participants were adults between the ages of 19 and 52. Mean age of the autism group was ∼7 years older than the neurotypical group and although this was a statistically significant difference, all participants were over 18 years and age was not related to aperiodic exponent at any dose in this study (all p’s > 0.05) and so was unlikely to contribute to the pattern of results reported.

**Table 1.** Demographic data

| Measure | N (M/ F) | Age | FSIQ | VIQ | PIQ | AQ |
| --- | --- | --- | --- | --- | --- | --- |
| Non-autistic | 8/13 | 29.00 (1.60) | 121.21 (2.06) | 119.37<br>(2.73) | 118.63 (1.92) | 15.78 (1.60) |
| Autistic | 9/6 | 37.27 (2.58) | 119.00 (2.43) | 117.45<br>(2.98) | 116.64 (2.69) | 35.87 (1.81) |
| *P-value* | .883 | .007* | .506 | .656 | .544 | < .001* |

Note: Demographic variables for each group are given including number (N) of males and females (M/F). Means (standard errors) are given for age, full-scale intelligence quotient (FSIQ), verbal intelligence quotient (VIQ), performance intelligence quotient (PIQ) and autism quotient. A chi-square test was used to compare the ratio of males to females between groups; between group t-tests were conducted to compare age, IQ, and AQ (bottom row)

### EEG data acquisition

Scalp EEG signals were collected using a 64-channel standard actiCAP (EASYCAP GmbH) with a sampling rate of 5 kHz and amplified by a BrainAmp amplifier (Brain Products GmbH). Electrode placements followed the international 10-20 system. Impedances between the scalp and electrodes were kept below 15 kilohms. Data were recorded relative to an FCz reference and a ground electrode was located at FPz. Participants took part in a resting-state protocol at the start of the EEG session, seated in a darkened room in front of a stimulus computer. The resting-state paradigm consisted of 6 x 1 minute trials that were either “eyes-open” or “eyes-closed”, presented alternately, the order of which was counterbalanced across participants. During “eyes-closed” trials, participants were asked to close their eyes; during “eyes-open” trials, participants were asked to look at a 1-minute sand-timer.

#### EEG data pre-processing

Raw data files were down sampled to 1000Hz. MATLAB 2016(a) and EEGLAB v14.1.2 were used for data pre-processing. Filters were adapted to suit the sampling rate using a Hamming windowed FIR filter, applied sequentially, and starting with the high pass filter of 2Hz passband, with a 1Hz cut-off at −6dB. A low-pass filter with a passband edge of 26.68Hz and a 30Hz cut-off was used. Areas of significantly noisy data were removed using manual continuous data rejection. Noisy and flat channels were identified manually and removed. Bad channels were then interpolated and the data was re-referenced to an average of all electrodes. Principal component analysis was used to reduce the dimensionality of the components down to the number of independent sources (i.e. non-interpolated channels). Independent Component Analysis was conducted using the AMICA algorithm. After ICA decomposition, eye blinks and saccadic ICA components were identified manually based on the time series and the topography of the components and removed from the data. Data were then segmented into two separate files, one containing three eyes open trials and one containing three eyes closed trials. All files were then converted from .set to .mat.

Power spectral densities (PSD’s) were computed in python 3.8 using neuro digital signal processing (NeuroDSP 2.1.0) toolbox. The channels for analysis were chosen based on topographic maps of aperiodic 1/f exponent values across groups, at placebo (Fig 1A, B) which illustrate that aperiodic exponents were maximal (i.e. largest) along the midline but less so at Fz in eyes closed trials and Oz. Therefore, channels used were Cz, CPz, Pz and POz. This is also in concordance with the literature that exists in terms of adult, aperiodic topographies (Jacob et al., 2021; Wang et al., 2022). PSD’s were computed over a frequency range of 3-28Hz using Welch’s method, taking the mean over windows. We used a segment length of 2000 samples with 50% overlap. To extract aperiodic exponents, the fitting oscillations and one over f (FOOOF 1.0.0) model was used (Donoghue, Haller, et al., 2020). Parameters were chosen based on extensive data quality checks and agreed upon by two experienced EEG researchers, these were: peak_width_limits=[1, 12], min_peak_height = 0.15, max_n_peaks=6, peak_threshold=2, aperiodic_mode=’fixed’.

**Figure 1.**
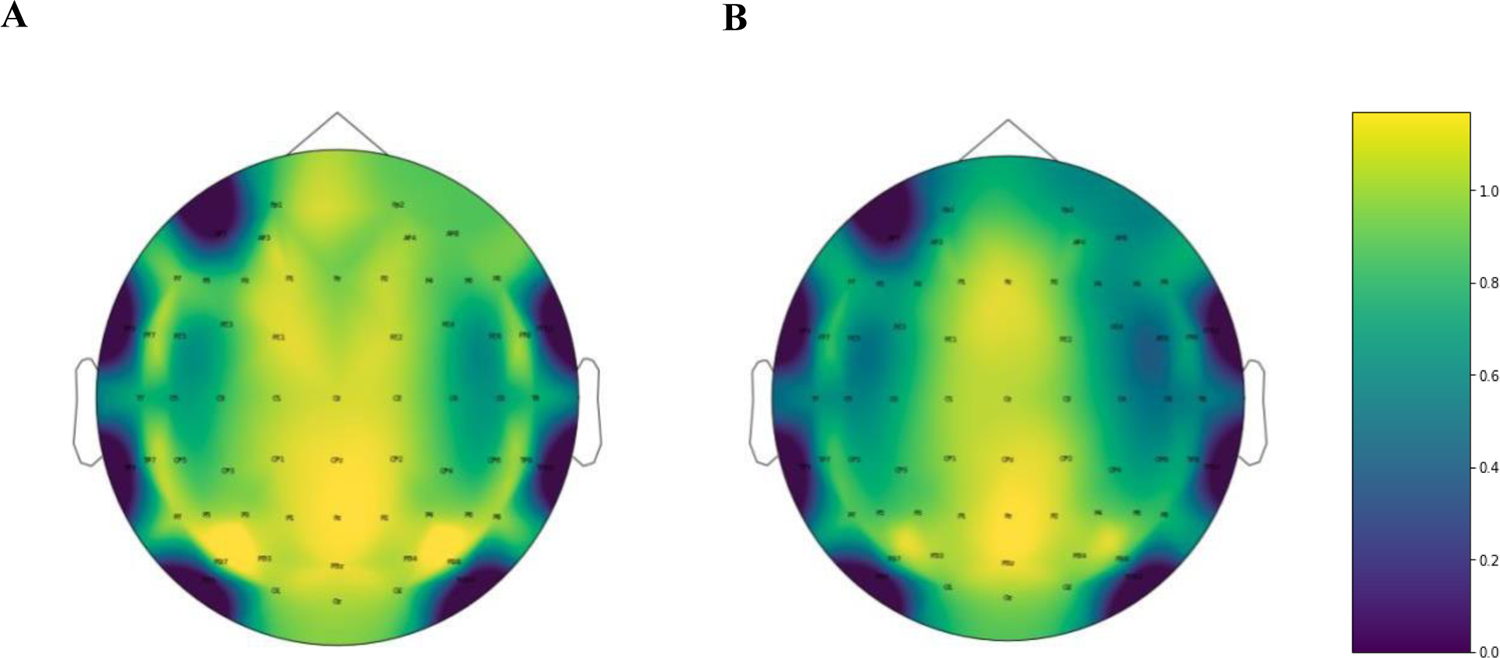
Topographic maps of aperiodic exponents across the scalp. Scalp distribution of aperiodic fit parameters indicates a midline, central to posterior parietal maximum for the aperiodic exponent in eyes closed trials (A) and a midline maximum for the aperiodic exponent in eyes open trials (B).

## Results

To address our hypotheses, we first analysed the data of participants who had aperiodic 1/f exponents for all three visits i.e. ‘three-visit analyses’. Therefore, we conducted 3×2 full-factorial mixed ANOVAs with drug dose as the within participants independent variable (placebo, 15 mg of arbaclofen, 30 mg of arbaclofen) and group (autism or neurotypical) as the between participants independent variable. We did this separately for eyes open and eyes closed trials. Normality checks were carried out on the residuals which were approximately normally distributed. Greenhouse-Geisser correction was used. Due to challenges in data collection (see methods section), not all subjects had data for each drug dose. The way mixed ANOVA models deal with missing data means that one missed visit eliminates data from all other visits of that participant. Therefore, to maximize power post-hoc, main effects from the omnibus analysis were followed up with separate 2×2 full-factorial mixed ANOVAs for participants with aperiodic 1/f exponents at placebo and 15 mg of arbaclofen and then for participants who had aperiodic exponents at placebo and 30 mg of arbaclofen; i.e. ‘two-visit analyses’. Normality checks were carried out on the residuals which were approximately normally distributed. Effect sizes were quantified using partial eta squared (η^2^) where η^2^ = 0.01 indicates a small effect, η^2^ = 0.06 indicates a medium effect and η^2^ = 0.14 indicates a large effect.

### Three visit analyses

Overall, 25 people were included in this analysis; 13 non-autistic individuals and 12 autistic individuals. Descriptive statistics are displayed in table 2. For eyes closed trials (see Fig 2A), results of the 3×2 mixed ANOVA confirmed the prediction that aperiodic 1/f exponents would increase (steepen) with increasing dose of arbaclofen; there was a main effect of drug dose [F(1.484, 34.139) = 9.444, p <.001, η^2^ = .291, large]. There was no main effect of group [F(1, 23) = .001, *p* = .972, partial eta squared = .000, small], and the drug x group interaction did not reach significance, however effect size was medium here [F(2, 46) = 3.396, p = .058, partial eta squared = .129, medium].

**Figure 2.**
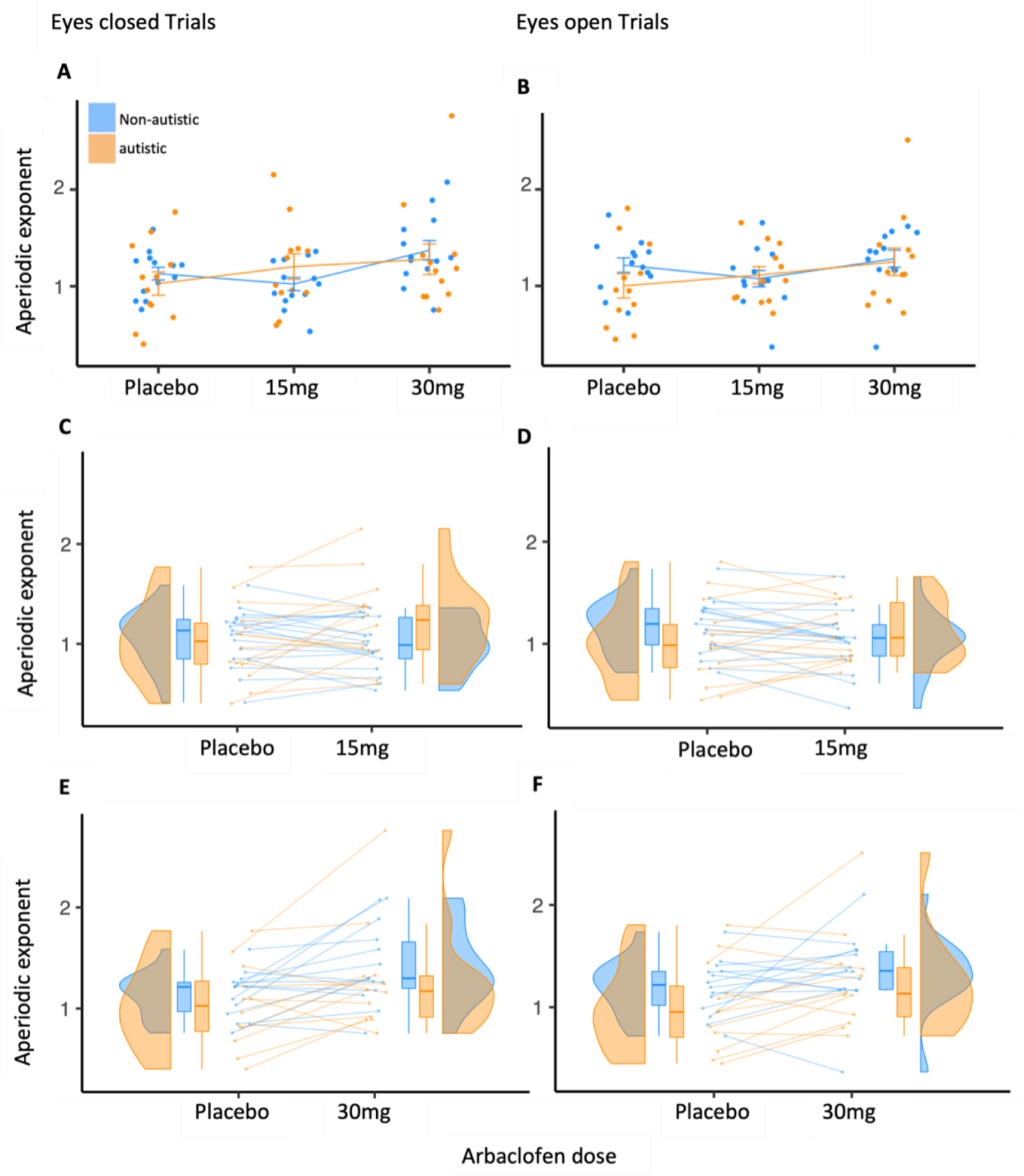
Aperiodic responses to drug dose in autistic and non-autistic people. Aperiodic 1/f exponents extracted during eyes-closed trials (**A**) Participants with all 3 doses, N= 25; (**B**) Participants with a placebo and low (15 mg) dose of arbaclofen, N = 31; (**C**) Participants with a placebo and high (30 mg) dose of arbaclofen, N = 26. Aperiodic 1/f exponents extracted during eyes-open trials (**D**) Participants with all 3 doses, N= 25; (**E**) Participants with a placebo and low (15 mg) dose of arbaclofen, N = 31; (**F**) Participants with a placebo and high (30 mg) dose of arbaclofen, N = 26. X-axes represent arbaclofen dose, y-axes represent aperiodic 1/f exponent. Error bars in (A) and (D) represent SEM

**Table 2.** Aperiodic 1/f exponents of autistic and non-autistic participants, at placebo, 15 mg of arbaclofen and 30 mg of arbaclofen

| Group | Trial | Drug dose |  |  |  |
| --- | --- | --- | --- | --- | --- |
|  |  | Placebo | 15 mg of arbaclofen | 30 mg of arbaclofen | Total |
| Non-autistic<br>(n = 13) | Closed | 1.131 (0.237) | 1.023 (0.244) | 1.372 (0.362) | 1.176 (0.316) |
|  | Open | 1.216 (0.270) | 1.076 (0.305) | 1.283 (0.320) | 1.192 (0.304) |
| Autistic<br>(n = 12) | Closed | 1.028 (0.418) | 1.204 (0.456) | 1.280 (0.547) | 1.171 (0.475) |
|  | Open | 1.002 (0.435) | 1.113 (0.303) | 1.250 (0.490) | 1.122 (0.418) |
| Total<br>(n = 25) | Closed | 1.082 (0.333) | 1.110 (0.365) | 1.328 (0.453) |  |
|  | Open | 1.113 (0.367) | 1.094 (0.298) | 1.267 (0.402) |  |

*Note.* Means (and standard deviations) of aperiodic 1/f exponents for each group at placebo, 15 mg of arbaclofen and 30 mg of arbaclofen, totals are given for each group across drug doses (rightmost column) and for each drug dose, across groups (bottom row)

For eyes open trials (See Fig 2B), the 3×2 mixed ANOVA confirmed a statistically significant main effect of drug dose [F(2, 46) = 7.276, *p* =.002, η^2^ = .240, large]. There was no main effect of group [F(1, 23) =.281, *p* = .601, η^2^ = .012, small]. Again the drug x group interaction did not reach significance but the effect size here was medium [F(2, 46) = 3.379, p = .060, partial eta squared = .128, medium].

### Two visit analyses

#### Placebo to 15 mg of arbaclofen

Overall 31 participants were included in this analysis; 17 non-autistic people and 14 autistic people. Descriptive statistics are displayed in table 3. Full statistics are presented in supplementary table 1. For both eyes closed and eyes open trials (see Fig 2 C, D), results of the 2×2 ANOVA comparing aperiodic 1/f exponents at placebo to aperiodic 1/f exponents at 15 mg of arbaclofen indicated there was no main effect of dose or group. However, there was a significant group by dose interaction (*p’s* < .05, η^2^*’s* = large). In eyes closed trials, in the neurotypical group, aperiodic 1/f exponents did not significantly differ between placebo and 15 mg of arbaclofen (*p* = .237, η^2^ = .048, small); whereas in the autism group, aperiodic 1/f exponents significantly increased from placebo to 15 mg of arbaclofen (*p* = .003, η^2^ = .262, large). Further, in eyes open trials, in the neurotypical group, aperiodic 1/f exponents significantly decreased from placebo compared to 15 mg of arbaclofen (*p* = .046, η^2^ = .130, medium). By contrast, in the autism group, planned comparisons showed that aperiodic 1/f exponents significantly increased from placebo to 15 mg (*p* = .009, η^2^ = .215, large).

**Table 3.** Aperiodic 1/f exponents of autistic and non-autistic participants, at placebo and 15 mg of arbaclofen

| Group | Trial | Drug dose |  |  |
| --- | --- | --- | --- | --- |
|  |  | Placebo | 15 mg of arbaclofen | Total |
| Non-autistic<br>(n = 17) | Closed | 1.063 (0.291) | 0.992 (0.261) | 1.027 (0.275) |
|  | Open | 1.164 (0.273) | 1.032 (0.303) | 1.098 (0.292) |
| Autistic<br>(n = 14) | Closed | 1.020 (0.391) | 1.223 (0.430) | 1.124 (0.416) |
|  | Open | 1.017 (0.403) | 1.125 (0.298) | 1.071 (0.353) |
| Total<br>(n = 31) | Closed | 1.044 (0.335) | 1.098 (0.361) |  |
|  | Open | 1.098 (0.341) | 1.074 (0.299) |  |

*Note.* Means (and standard deviations) of aperiodic 1/f exponents for each group at each drug dose for placebo and 15 mg of arbaclofen. Means are also included for each drug dose, across groups (bottom row), and for each group, across drug dose (rightmost column)

#### Placebo to 30 mg of arbaclofen

Overall 26 people were included in this analysis, 14 non-autistic people and 12 autistic people. Descriptive statistics are displayed in table 4. Full statistics are presented in supplementary table 2. For both eyes closed and eyes open trials (see Fig 2E, F), results of the 2×2 ANOVA comparing aperiodic 1/f exponents at placebo to aperiodic 1/f exponents at 30 mg of arbaclofen revealed a significant main effect of drug (*p’s* < .05, η^2^*’s* = large) whereby aperiodic 1/f exponents at 30 mg of arbaclofen were significantly larger (steeper), across both groups, compared to those at placebo. There was no main effect of group, or drug x group interaction. In the neurotypical group, aperiodic 1/f exponents significantly increased from placebo compared to 30 mg (closed-*p* = .013, η^2^ = .231, large; open-p = .032, η^2^ = .177, large). Similarly, in eyes closed trials, in the autism group aperiodic 1/f exponents significantly increased from placebo to 30 mg (*p* = .039, η^2^ = .166, large). In eyes open trials, aperiodic 1/f exponents increased from placebo to 30 mg in autistic individuals, but this was not significant (p = .160, η^2^ = .081, medium).

**Table 4.**
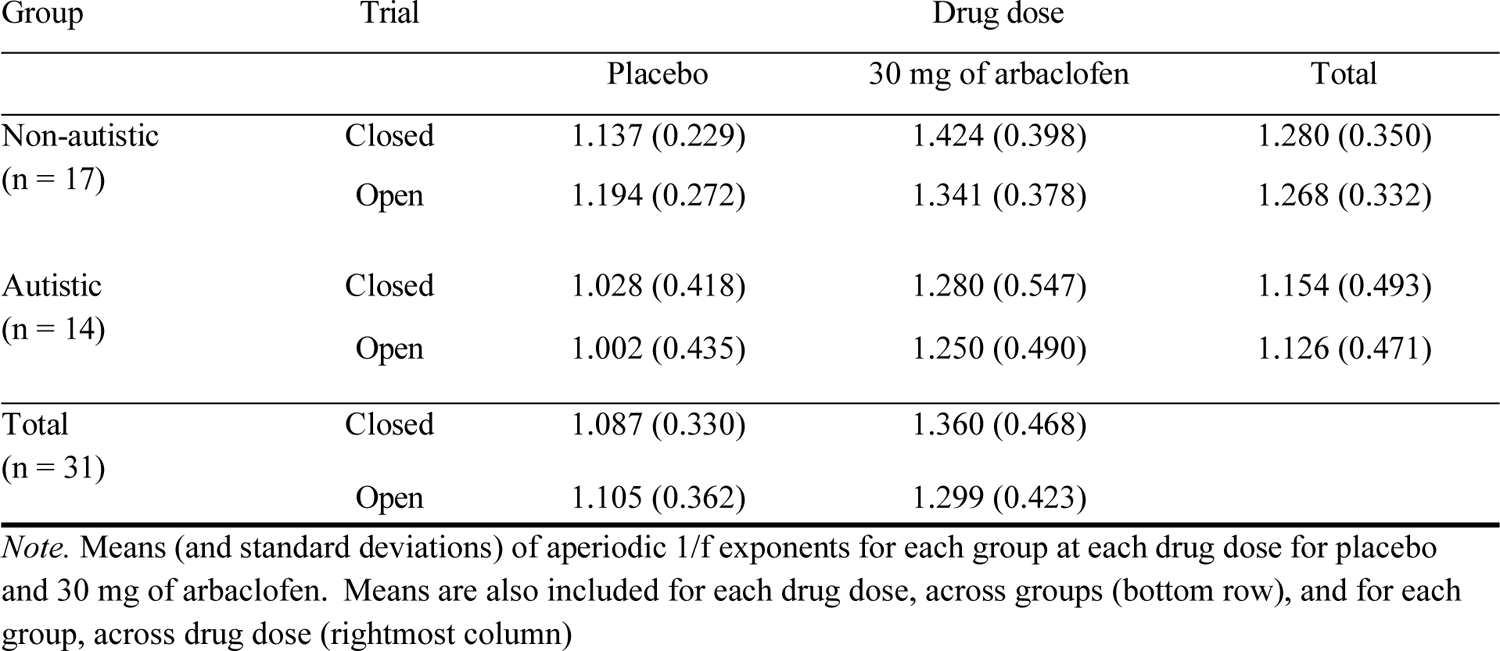
Aperiodic 1/f exponents of autistic and non-autistic participants, at placebo and 30 mg of arbaclofen

## Discussion

Here we report, for the first time, that in humans the aperiodic 1/f exponent of the EEG power spectrum is responsive to E-I challenge with the GABA agonist arbaclofen; and that, on average, responsivity differs between autistic and non-autistic people. Thus, we have confirmed the utility of the aperiodic 1/f exponent as an E-I sensitive metric in humans, and used it to reveal that (homeostatic) regulation of E-I dynamics are different in the neurodevelopmental condition autism.

In brief, across both groups, the highest dose of arbaclofen elicited a shift (increase) in aperiodic 1/f exponents, steepening the slope. As arbaclofen increases global inhibition by blocking glutamate release pre-synaptically, and acts as a GABA_B_ receptor agonist post-synaptically (de Groot & van Strien, 2018), these results are in line with animal findings of steeper aperiodic exponents with increased central inhibition (Gao, Peterson, Voytek, et al., 2017). These results add rigor to existing findings using broader-action pharmacological manipulations of E-I (Waschke et al., 2021). Furthermore, as EEG was recorded at rest, our results were not confounded by task effects (Molina et al., 2020; Waschke et al., 2021).

Overall there were no case-control group differences in aperiodic 1/f exponents between autistic and non-autistic people under placebo (baseline) or any drug condition. This appears to contradict theories predicting a relative increase in ‘excitation’ at baseline in autistic people (Sohal & Rubenstein, 2019) reports of smaller aperiodic 1/f exponents in preterm infants with increased likelihood of autism (Shuffrey et al., 2022) and in autistic children (with below-average IQ; Manyukhina et al., 2022). One explanation for this apparent discrepancy is simply that previous studies examined children whereas we recruited adults; indeed E-I pathways change with development (Dorrn et al., 2010) and aperiodic 1/f exponents decrease with age (Voytek et al., 2015). However, the heterogeneity of autism itself may also influence results (Dickinson et al., 2016; Loth, 2016; Loth et al., 2021; Mottron & Bzdok, 2020).

In addition, the aperiodic 1/f exponent is a dynamic measure. In the placebo condition, at rest, we likely capture the E-I system in a baseline state of flux. Introducing a challenge to disrupt the system with arbaclofen may make very different demands on autistic and non-autistic E-I systems. In line with this, we observed a different drug response between groups at 15 mg of arbaclofen; on average, aperiodic 1/f exponents in the autism group were steeper in response to 15 mg of arbaclofen (both eyes closed and eyes open) but showed no change (eyes closed) or flattened (eyes open) in the neurotypical group. One explanation for this could be that the compensatory mechanisms in autistic adults which serve to maintain overall E-I homeostasis in the baseline condition can be over-ridden at a low dose because their dynamic regulation is different (Sohal & Rubenstein, 2019).

That is, E-I circuits are not static, they constantly adjust in response to the environment. Sensory inputs destabilise E-I circuits so that tightly controlled E-I dynamics are established early in development to offset destabilisation (Dorrn et al., 2010), avoid run away or silent activity (Litwin-Kumar & Doiron, 2012), and fine-tune sensory response (Heiss et al., 2008; Mariño et al., 2005; Poo & Isaacson, 2009; Tao & Poo, 2005; Wehr & Zador, 2003). Maintenance of E-I homeostasis is achieved by a set of synaptic plasticity mechanisms such as synaptic scaling; whereby E or I synaptic strength is adjusted up or down to stabilise firing rate (Turrigiano, 2011, 2012; Turrigiano, 1999; Turrigiano & Nelson, 2004). Therefore, in challenging the system with arbaclofen, we may have exposed the result of underlying homeostatic E-I differences that occur on fast timescales. Due to the excellent temporal resolution of EEG, these dynamic differences in E-I signalling were captured by the aperiodic 1/f exponent.

### Application of the aperiodic 1/f exponent

The utility of the aperiodic 1/f exponent, as a metric of E-I, for psychiatric research has been questioned (Bruining et al., 2020) as there is no absolute value to indicate whether someone has an E or I dominant regime. However, at rest, “somehow the unstable stuff of which we are composed has learned the trick of maintaining stability” (Cannon, 1932). We would argue that the utility of this metric lies in its ability capture a *shift* from stability, i.e. brain dynamics. We have shown that, in combination with single dose drug challenge designs which in our lab we have termed ‘shiftability studies’, the aperiodic 1/f exponent can expose dose-dependent differences in E-I flux. Indeed, whilst this study does not consider the clinical efficacy of arbaclofen, it nonetheless has important implications for the development of pharmacological support options for autistic individuals. The next steps might include assessing if aperiodic shift elicited by a candidate compound could be used to predict an individual’s clinical response to drug.

### Limitations

Arbaclofen is not without side effects which can impact on data collection. In our study these were restricted to its known side effects (particularly dizziness and nausea). We adapted the study design to minimize the chances of discomfort; if participant had uncomfortable side effects at a low dose, they did not attend the high dose visit (see methods). To accommodate this sample size variation, results were interpreted with their corresponding effect sizes, which were all medium to large. Furthermore, the current sample size far exceeds those used in the preclinical results this study sought to replicate (Gao, Peterson, Voytek, et al., 2017). Indeed, as this was a PoC study, designed to assess translation of animal evidence for 1/f as a measure of E-I dynamics to humans: to that end, the results from this sample support our aim.

In conclusion, here we provide PoC evidence that the aperiodic 1/f exponent is responsive to E-I pharmacological challenge in humans thus bridging an oft-overlooked gap between preclinical and human studies. Critically these results were achieved with a non-invasive, cheap method in combination with a completely passive task that is not cognitively demanding. This opens up multiple opportunities for future research. Within autism for example, there may be scope to extend this tool to ensure those with intellectual disability are included in research into E-I. Our PoC shows that, in humans, the aperiodic 1/f exponent captures individual E-I dynamics and opens the potential for this metric to be adopted in research which target E-I homeostasis for potential pharmacological support options for neurodevelopmental and/or other psychiatric conditions with E-I alterations. Because the aperiodic 1/f signal is observed at multiple different scales (i.e. Local Field Potential, Electrocorticography and EEG) and across species, it could help bridge the translational gap which so often separates preclinical and clinical neuroscience research.

## Data Availability

All data produced in the present study are available upon reasonable request to the authors

