## Supplemental Information for "The dynamically neurodiverse human brain: Measuring excitatory-inhibitory dynamics and identifying homeostatic differences in autistic and non-autistic people"

### Supplementary Information

#### Supplementary Table 1

*Placebo to 15 mg of arbaclofen*

| | Main effect | F-statistic | p-value | Effect size |
|---|---|---|---|---|
| Closed | Drug | 2.444 | .129 | .078, medium |
| | Group | .688 | .414 | .023, small |
| | Drug x Group | 10.142 | .003* | .259, large |
| Open | Drug | .122 | .730 | .004, small |
| | Group | .061 | .806 | .002, small |
| | Drug x Group | 11.816 | .002* | .289, large |

Note: F-statistic , p-value and effect sizes are presented for each main effect for each trial-type

#### Supplementary Table 2

*Placebo to 30 mg of arbaclofen*

| | Main effect | F-statistic | p-value | Effect size |
|---|---|---|---|---|
| Closed | Drug | 11.756 | .002* | .329, large |
| | Group | .817 | .375 | .033, small |
| | Drug x Group | .048 | .828 | .002, small |
| Open | Drug | 7.044 | .014* | .277, large |
| | Group | 1.071 | .311 | .043, small |
| | Drug x Group | .467 | .501 | .019, small |

Note: F-statistic , p-value and effect sizes are presented for each main effect for each trial-type
